# Complement dysregulation is a predictive and therapeutically amenable feature of long COVID

**DOI:** 10.1101/2023.10.26.23297597

**Authors:** Kirsten Baillie, Helen E Davies, Samuel B K Keat, Kristin Ladell, Kelly L Miners, Samantha A Jones, Ermioni Mellou, Erik J M Toonen, David A Price, B Paul Morgan, Wioleta M Zelek

**Affiliations:** Division of Infection and Immunity, Cardiff University School of Medicine, University Hospital of Wales, Heath Park, Cardiff CF14 4XN, UK; Department of Respiratory Medicine, University Hospital of Wales, Llandough, Penarth CF64 2XX, UK; R&D Department, Hycult Biotechnology, Frontstraat 2A, 5405 PB, Uden, The Netherlands; Systems Immunity Research Institute, Cardiff University School of Medicine, University Hospital of Wales, Heath Park, Cardiff CF14 4XN, UK

## Abstract

**Background:** Long COVID encompasses a heterogeneous set of ongoing symptoms that affect many individuals after recovery from infection with severe acute respiratory syndrome coronavirus 2 (SARS-CoV-2). The underlying biological mechanisms nonetheless remain obscure, precluding accurate diagnosis and effective intervention. Complement dysregulation is a hallmark of acute COVID-19 but has not been investigated as a potential determinant of long COVID.

**Methods:** We quantified a series of complement proteins, including markers of activation and regulation, in plasma samples from healthy convalescent individuals with a confirmed history of infection with SARS-CoV-2 and age/ethnicity/gender/infection/vaccine-matched patients with long COVID.

**Findings:** Markers of classical (C1s-C1INH complex), alternative (Ba, iC3b), and terminal pathway (C5a, TCC) activation were significantly elevated in patients with long COVID. These markers in combination had a receiver operating characteristic predictive power of 0.794. Other complement proteins and regulators were also quantitatively different between healthy convalescent individuals and patients with long COVID. Generalized linear modeling further revealed that a clinically tractable combination of just four of these markers, namely the activation fragments iC3b, TCC, Ba, and C5a, had a predictive power of 0.785.

**Conclusions:** These findings suggest that complement biomarkers could facilitate the diagnosis of long COVID and further suggest that currently available inhibitors of complement activation could be used to treat long COVID.

**Funding:** This work was funded by the National Institute for Health Research (COV-LT2-0041), the PolyBio Research Foundation, and the UK Dementia Research Institute.

## INTRODUCTION

The severe acute respiratory syndrome coronavirus 2 (SARS-CoV-2) pandemic left a legacy of chronic illness in a large proportion of survivors of acute coronavirus disease 19 (COVID-19). Encapsulated under the umbrella term “post-acute sequelae of SARS-CoV-2”, the presence of new or ongoing symptoms more than 12 weeks after the acute infection is most commonly known as long COVID.^1–3^ The spectrum of disease is extensive and variable. Common symptoms include cognitive blunting, also called “brain fog”, chest pain, dyspnoea, fatigue, and sensory dysregulation, which often have a substantial impact on daily activities and quality of life, akin to myalgic encephalomyelitis/chronic fatigue syndrome.^4^ A recent systematic review concluded that 45% of individuals experience diverse and unresolved symptoms 4 months after infection with SARS-CoV-2, irrespective of initial disease severity.^5^ Chronic disease is also common. For example, a recent national survey (https://www.ons.gov.uk/peoplepopulationandcommunity/healthandsocialcare/conditionsanddiseases/articles/coronaviruscovid19latestinsights/infections#long-covid) found that 1.9 million people (2.9% of the population) reported symptoms compatible with long COVID in the UK, with an estimated 41% of affected individuals experiencing ongoing ill health for at least 2 years as of March 2023.

Despite the profound burden of suffering and socioeconomic consequences of long COVID, it remains unclear how chronic illness develops and persists after infection with SARS-CoV-2. Several mechanisms have been proposed to account for the pathogenesis of long COVID, including viral persistence, endothelial dysfunction, coagulation defects, and immune dysregulation.^6^ Persistent inflammation, signposted by elevated blood concentrations of C-reactive protein and proinflammatory cytokines, has also been reported in people with long COVID.^7,8^ The underlying causes of this inflammatory process nonetheless remain obscure. Dysregulation of the complement cascade has been implicated as a driver of inflammation in many diseases, including acute COVID-19.^9–12^ Indeed, the complement system is markedly dysregulated in severe acute COVID-19, and biomarkers spanning all activation pathways predict disease outcome.^10,13–17^ On the basis of these observations, we hypothesized that complement dysregulation could play a key role in the pathogenesis of long COVID.

To evaluate this hypothesis, we conducted an extensive analysis of the complement system in plasma samples obtained from a large cohort of age/ethnicity/gender/infection/vaccine-matched healthy convalescent individuals and non-hospitalized patients with long COVID. Activation products demarcating the classical, alternative, and terminal complement pathways were significantly elevated in patients with long COVID relative to healthy convalescent individuals after recovery from infection with SARS-CoV-2. Plasma concentrations of some complement components and regulators also differed significantly between healthy convalescent individuals and patients with long COVID. Moreover, various combinations of these analytes, including minimal panels with clinical applicability, were highly predictive of disease. These findings implicate complement dysregulation as a driver of inflammation and provide a novel set of biomarkers that could aid the diagnosis and guide the treatment of long COVID.

## RESULTS

### Complement activation products are elevated in patients with long COVID

The role of complement dysregulation as a potential determinant of long COVID symptomatology has not been investigated previously. To address this knowledge gap, we first quantified six markers of complement activation, including classical, lectin, alternative, and terminal pathway products, in plasma samples obtained from healthy convalescent individuals (controls, *n* = 79) and patients with long COVID (cases, *n* = 166). All participants had a clearly defined episode of acute COVID-19 confirmed via molecular evidence of infection with SARS-CoV-2. Groups were matched for age (cases, median = 47 years; controls, median = 45 years), ethnicity (cases, white = 88.0%; controls, white = 84.8%), gender (cases, female = 76.5%; controls, female = 78.5%), infection wave (cases, 54.8% infected more than 2 years before sample acquisition; controls, 46.8% infected more than 2 years before sample acquisition), and vaccination status (cases, median number of vaccinations before infection = 2; controls, median number of vaccinations before infection = 3), a parameter known to mitigate the risk of long COVID.^18^ Of note, obesity (BMI > 30) was significantly more common in cases versus controls (48.8% versus 34.6%, respectively; p = 0.042), and although employment status was comparable between groups pre-acute COVID-19, only 43.6% of cases remained in full-time work post-acute COVID-19 compared with 89.1% pre-acute COVID-19 (p < 0.00001). Cohort demographics, symptomatology, and other key features are summarized in **Supplementary Table 1**.

The C1s-C1INH complex, a product of classical pathway activation, was significantly elevated in cases versus controls (1.25 versus 1.09 µg/ml, respectively, p = 0.0089), whereas no such differences were observed for the mannose-associated serine protease 1 (MASP1)-C1INH complex, generated during lectin pathway activation (65.0 versus 56.8 ng/ml, respectively, p = 0.15) (**Figure 1A, B**). The fragments iC3b and Ba, which indicate alternative pathway activation, were also significantly elevated in cases versus controls (iC3b, 20.7 versus 14.4 µg/ml, respectively, p < 0.0001; Ba, 0.39 versus 0.22 µg/ml, respectively, p < 0.001) (**Figure 1 C, D**). In addition, C5a and the terminal complement complex (TCC), which demarcate terminal pathway activation, were both significantly elevated in cases versus controls, with the latter demonstrating a substantial increase (C5a, 7.45 versus 5.09 ng/ml, respectively, p < 0.02; TCC, 5.56 versus 3.55 µg/ml, respectively, p < 0.0001) (**Figure 1E, F**). Assay details are provided in **Supplementary Table 2**, and the results are summarized in **Table 1**.

**Figure 1.**
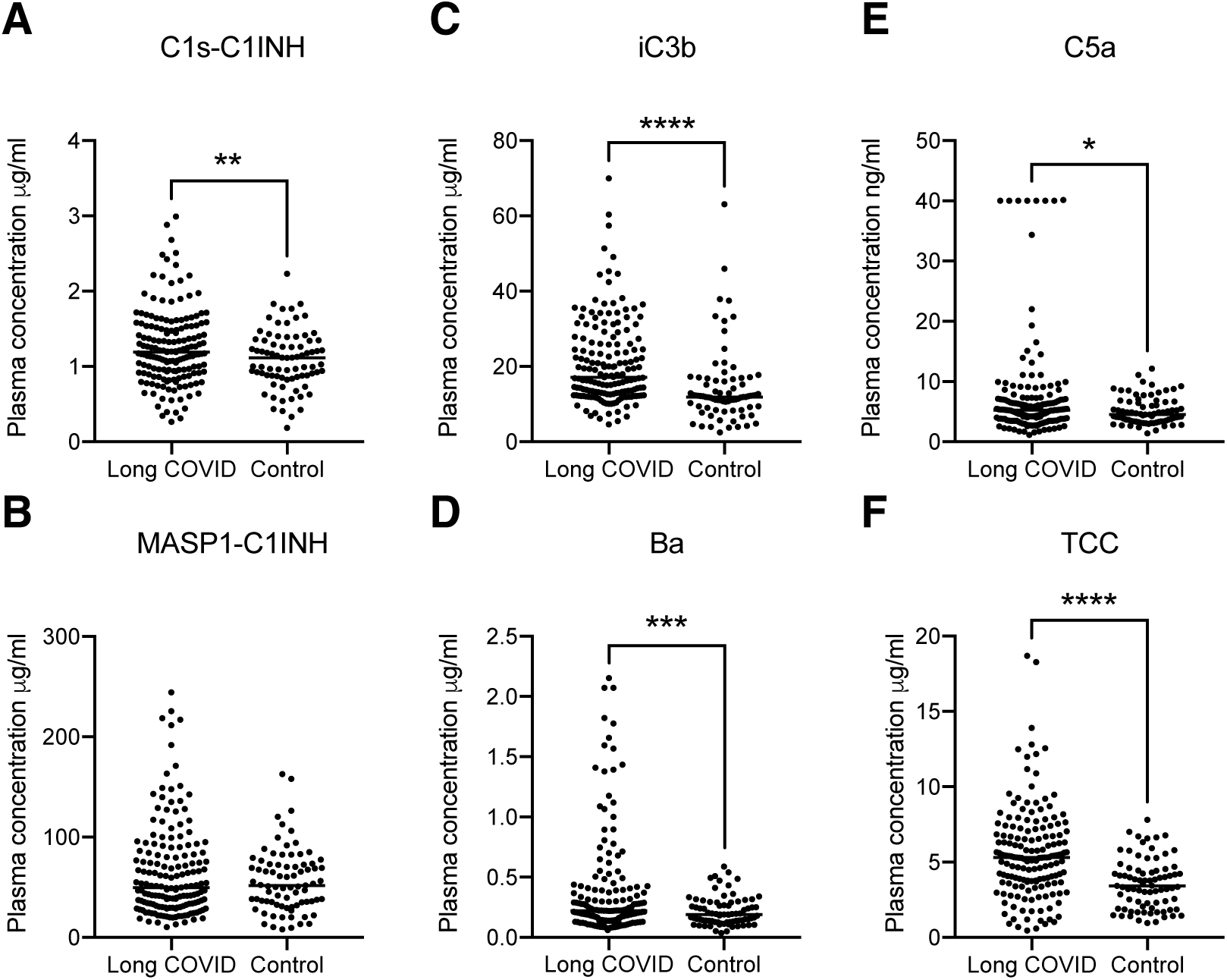
Plasma concentrations of complement activation products in healthy convalescent individuals and patients with long COVID. (**A**–**F**) Dot plots showing plasma concentrations of C1s-C1INH (**A**), MASP1-C1INH (**B**), iC3b (**C**), Ba (**D**), C5a (**E**), and TCC (**F**) in healthy convalescent individuals (*n* = 79) and patients with long COVID (*n* = 166). Horizontal bars represent mean values. *p < 0.05, **p < 0.01, ***p < 0.001, ****p < 0.0001 (unpaired t-test).

**Table 1.** Complement biomarker concentrations in healthy convalescent individuals and patients with long COVID.

|  | Units | Controls | Cases | P value |
| --- | --- | --- | --- | --- |
| **Activation products** |  |  |  |  |
| C1s-C1INH | µg/ml | 1.088 | 1.255 | **0.0089** |
| MASP1-C1INH | ng/ml | 56.75 | 65.03 | 0.154 |
| Ba | µg/ml | 0.2172 | 0.3871 | **0.0007** |
| iC3b | µg/ml | 14.36 | 20.72 | **<0.0001** |
| C5a | ng/ml | 5.090 | 7.454 | **0.0135** |
| TCC | µg/ml | 3.551 | 5.559 | **<0.0001** |
| **Components** |  |  |  |  |
| C1q | µg/ml | 130 | 109.2 | **0.0312** |
| C4 | µg/ml | 511.8 | 477.8 | 0.1341 |
| C3 | µg/ml | 830.2 | 893.3 | **0.0102** |
| FB | µg/ml | 279.7 | 268.7 | 0.5314 |
| C5 | µg/ml | 174.5 | 198.4 | **0.0037** |
| C9 | µg/ml | 83.2 | 92.52 | **0.0103** |
| **Regulators** |  |  |  |  |
| C1INH | µg/ml | 100.9 | 113.7 | **0.0011** |
| FH | µg/ml | 232.3 | 263.4 | 0.079 |
| FHR125 | µg/ml | 101.7 | 113.9 | 0.2866 |
| FHR4 | µg/ml | 3.879 | 3.883 | 0.9824 |
| sCR1 | ng/ml | 6.759 | 6.436 | 0.2141 |
| Properdin | µg/ml | 2.994 | 3.878 | **<0.0001** |
| FD | µg/ml | 0.6977 | 0.922 | **<0.0001** |
| FI | µg/ml | 54.17 | 56.53 | 0.2468 |
| Clusterin | µg/ml | 130.6 | 145.6 | **0.043** |

Mean values are shown. All units in µg/ml, except for MASP1-C1INH, C5a, and sCR1 (all ng/ml). P values were calculated using an unpaired t-test. Significant differences are highlighted in bold font. C1INH, C1 inhibitor; MASP1, mannose-associated serine protease 1; TCC, terminal complement complex; FB, factor B; FH, factor H; FHR125, FH-related proteins 1, 2, and 5; FHR4, FH-related protein 4; sCR1, soluble complement receptor 1; FD, factor D; FI, factor I.

### Complement components and regulators are altered in patients with long COVID

To extend these findings, we quantified a series of complement components and regulators in the same samples, again comparing healthy convalescent individuals (controls, *n* = 79) and patients with long COVID (cases, *n* = 166). C1q, the trigger for classical pathway activation, was significantly lower in cases versus controls (109.2 versus 130 µg/ml, respectively, p < 0.05), likely reflecting consumption (**Figure 2A**). In contrast, C3, C5, and C9 were all significantly elevated in cases versus controls (C3, 0.89 versus 0.83 mg/ml, respectively, p < 0.01; C5, 198.4 versus 174.5 µg/ml, respectively, p < 0.005; C9, 92.5 versus 83.2 µg/ml, respectively, p < 0.01) (**Figure 2 D–F**). All three of these proteins are positive acute phase reactants, likely explaining the increased concentrations in patients with long COVID. Levels of C4 and factor B (FB) were also higher in cases versus controls, albeit not significantly (**Figure 2 B, C**). Assay details are provided in **Supplementary Table 2**, and a data summary is provided in **Table 1**.

**Figure 2.**
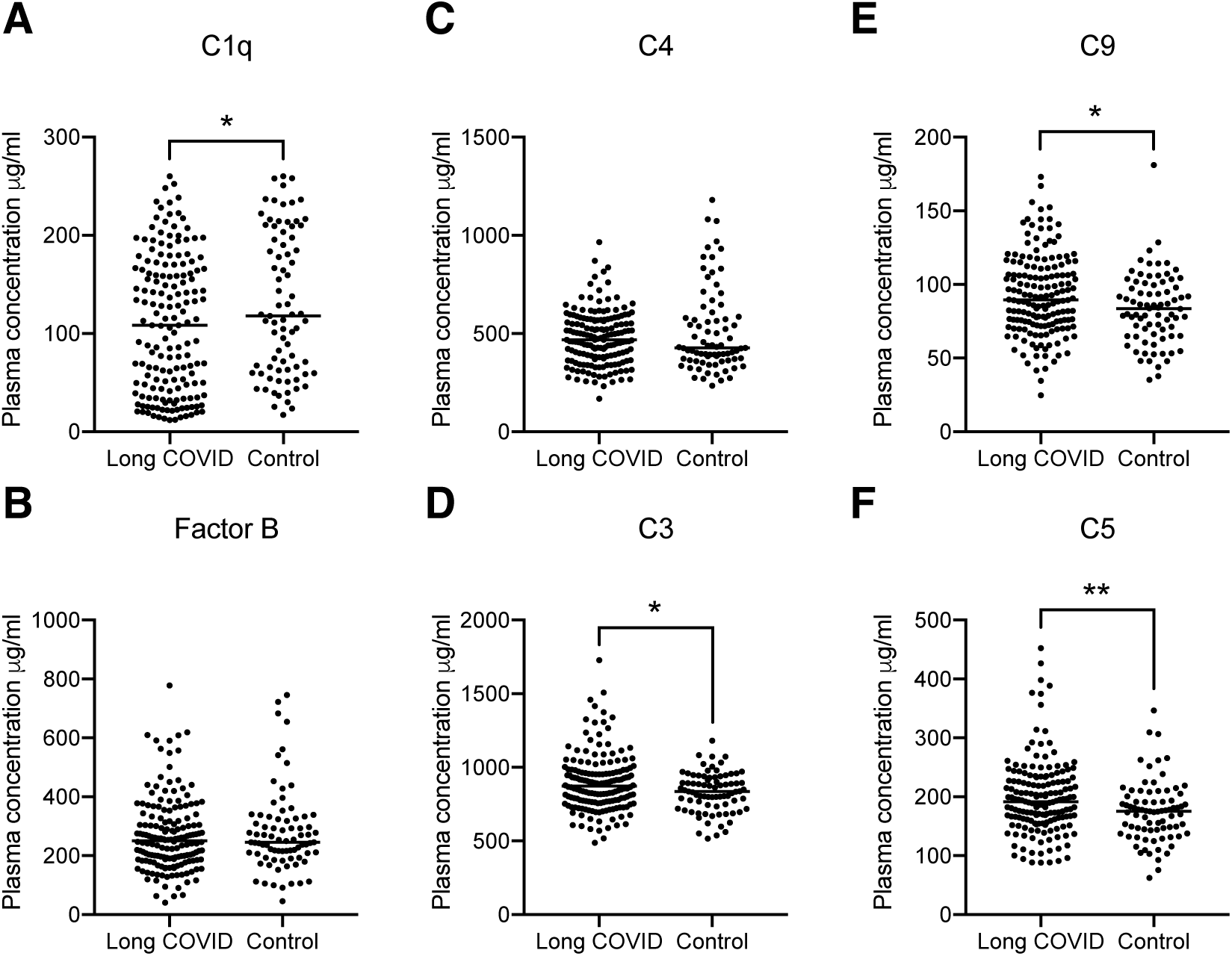
Plasma concentrations of complement components in healthy convalescent individuals and patients with long COVID. (**A**–**F**) Dot plots showing plasma concentrations of C1q (**A**), factor B (**B**), C4 (**C**), C3 (**D**), C9 (**E**), and C5 (**F**) in healthy convalescent individuals (*n* = 78–79) and patients with long COVID (*n* = 166). Horizontal bars represent mean values. *p < 0.05, **p < 0.01 (unpaired t-test).

Most of the complement regulators selected for measurement were also significantly elevated in cases versus controls, including C1INH, the key regulator of classical and lectin pathway activation (113.7 versus 100.9 µg/ml, respectively, p < 0.001) (**Figure 3A**), factor D (FD), factor H (FH), and properdin, which are involved in regulation of alternative pathway activation (FD, 0.92 versus 0.70 µg/ml, respectively, p < 0.0001; FH, 263.4 versus 232.3 µg/ml, respectively, p < 0.01; properdin, 3.88 versus 2.99 µg/ml, respectively, p < 0.0001) (**Figure 3 B–D**), and clusterin, a regulator of the terminal pathway (145.6 versus 130.6 µg/ml, respectively, p < 0.05) (**Figure 3E**). Plasma levels of the key alternative pathway regulator factor I (FI), the soluble form of complement receptor 1 (sCR1), and the FH-related (FHR) proteins (FHR4 and FHR125) were not significantly different between healthy convalescent individuals and patients with long COVID (**Figure 3F**, **Supplementary** Figure 1 **A**–**C**). Of note, there were also no differences in plasma haemolytic activity or anti-SARS-CoV-2 spike protein receptor-binding domain (RBD) antibody titers between healthy convalescent individuals and patients with long COVID (**Supplementary** Figure 1 **D**, **E**). Assay details are provided in **Supplementary Table 2**, and a data summary is provided in **Table 1**.

**Figure 3.**
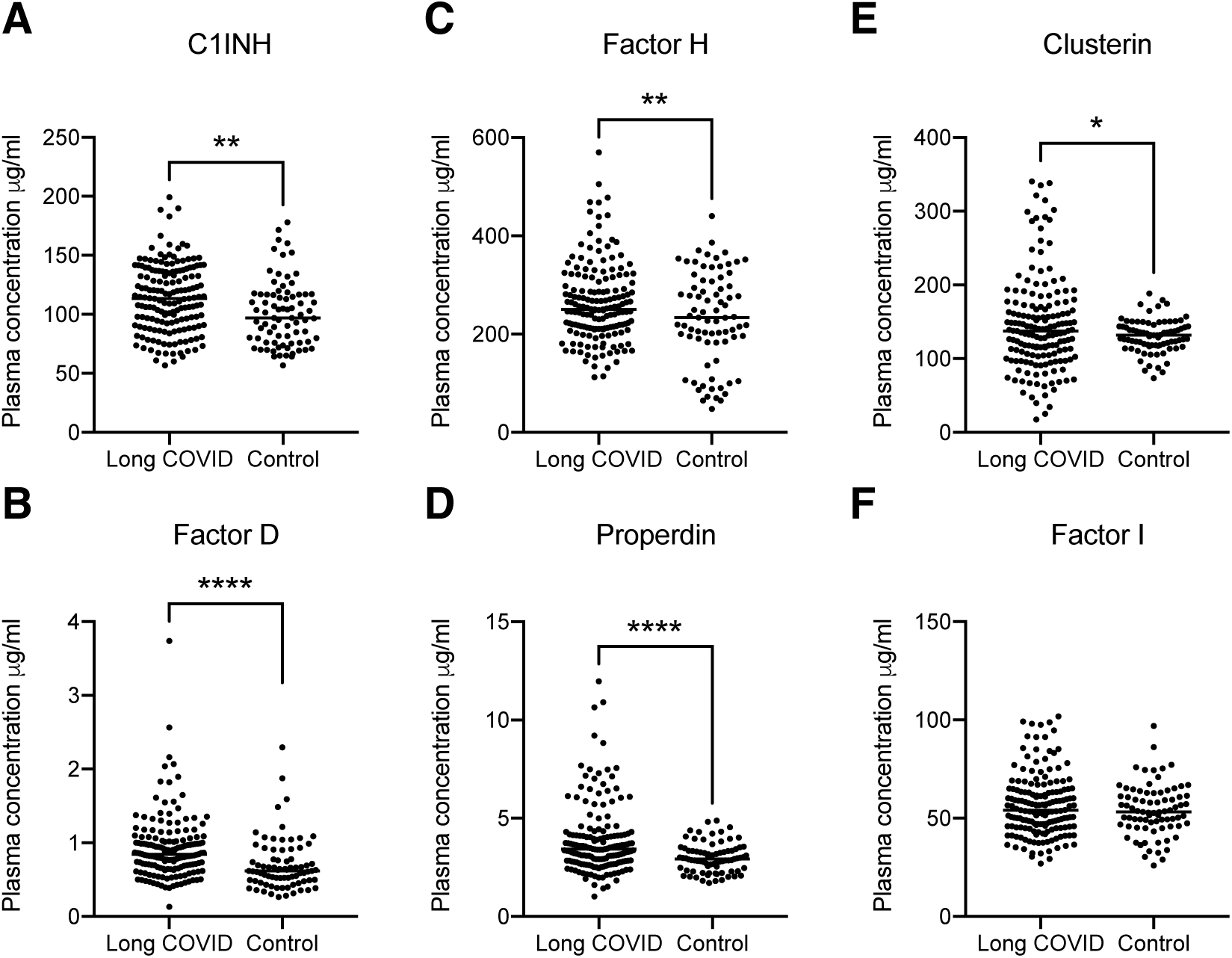
Plasma concentrations of complement regulators in healthy convalescent individuals and patients with long COVID. (**A**–**F**) Dot plots showing plasma concentrations of C1INH (**A**), factor D (**B**), factor H (**C**), properdin (**D**), clusterin (**E**), and factor I (**F**) in healthy convalescent individuals (*n* = 78–79) and patients with long COVID (*n* = 166). Horizontal bars represent mean values. *p < 0.05, **p < 0.01, ****p < 0.0001 (unpaired t-test).

### Complement biomarker sets identify patients with long COVID

Plasma concentration distributions for each complement protein measured in this study (*n* = 21) are shown for cases and controls as density plots in **Figure 4A** and as scatter plots versus age in **Figure 4B**. The associated statistics are shown in **Supplementary Table 3**. Pearson correlograms are shown in **Figure 5**. In receiver operating characteristic (ROC) analyses using multiple generalized linear models (GLMs), 9/21 complement proteins measured in our panels showed good predictive power (area under the curve [AUC] > 0.6), including three activation products (ranked iC3b, TCC, Ba) and six other markers (ranked C1INH, FD, C3, properdin, C9, FH) (**Figure 6A, B**). The most predictive single marker was C1INH, with an AUC of 0.746 (**Figure 6B**).

**Figure 4.**
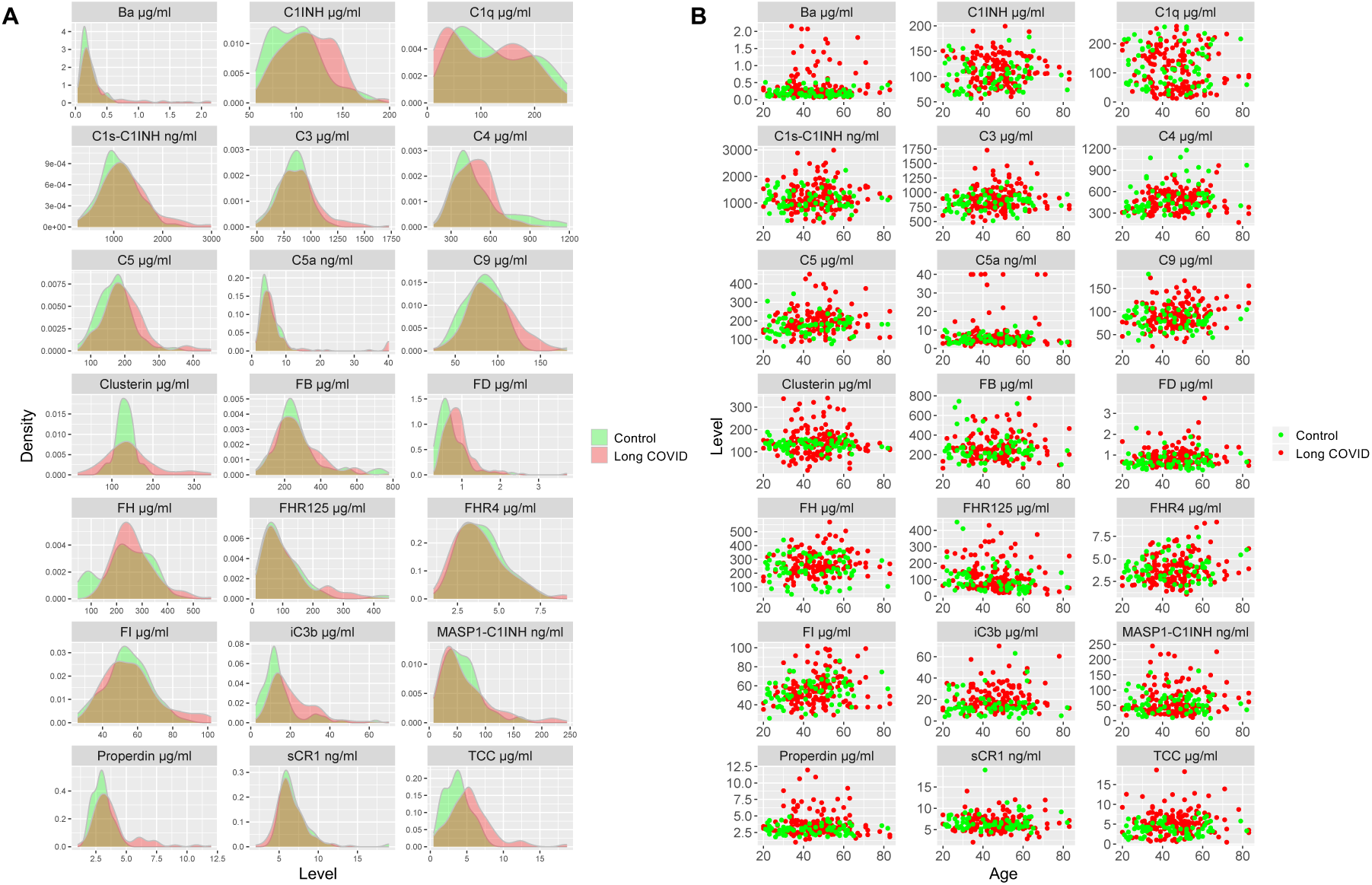
Plasma concentration distributions for complement proteins in healthy convalescent individuals and patients with long COVID. (**A**) Density plots showing the plasma concentration distribution for each complement analyte (*n* = 21) in healthy convalescent individuals (green, *n* = 78–79) and patients with long COVID (red, *n* = 166). (**B**) Scatter plots showing the plasma concentration distribution for each complement analyte (*n* = 21) versus age at inclusion in healthy convalescent individuals (green, *n* = 78–79) and patients with long COVID (red, *n* = 166).

**Figure 5.**
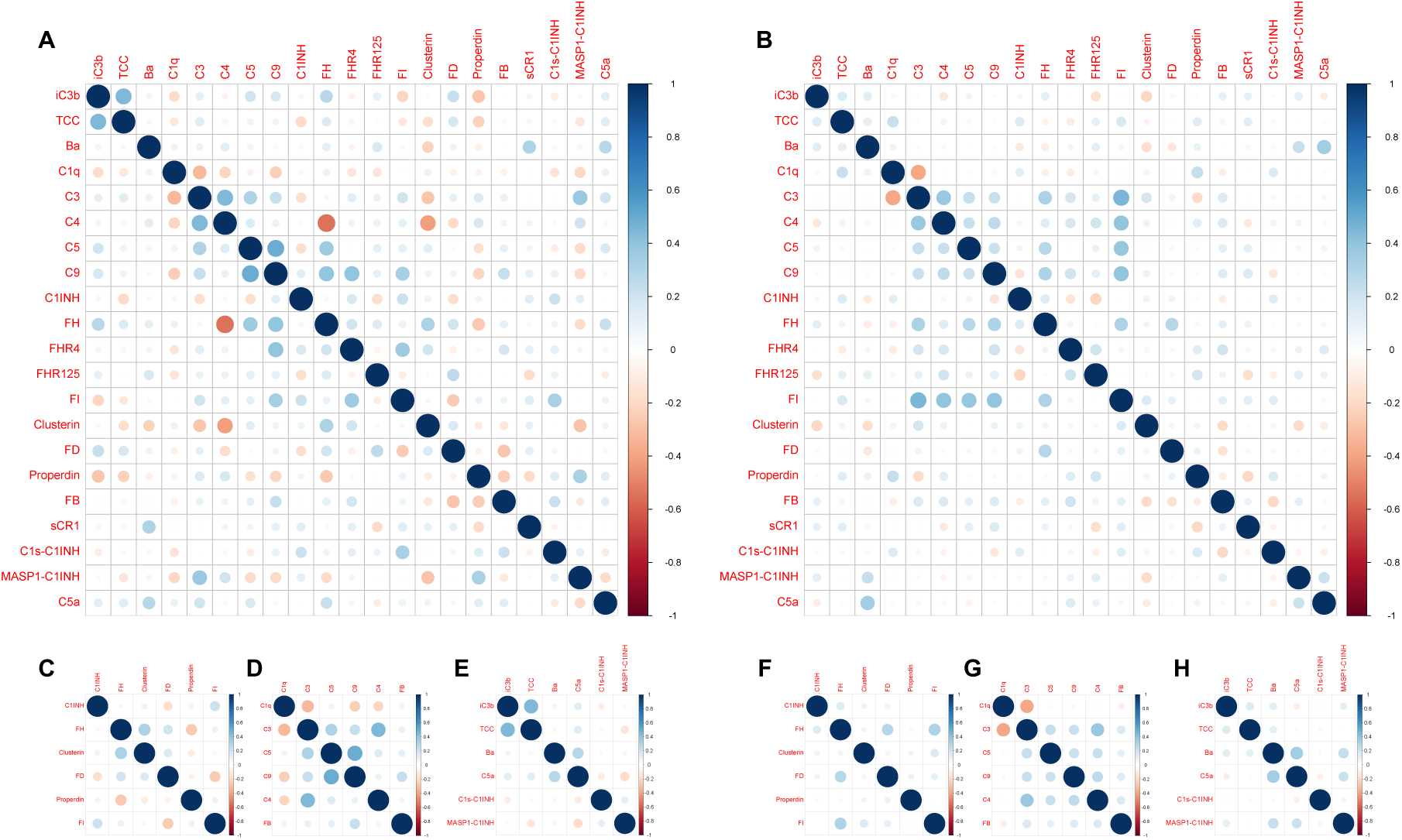
Complement analyte correlograms for healthy convalescent individuals and patients with long COVID. **(A–H)** Correlograms are shown for all complement analytes measured in plasma samples from healthy convalescent individuals (**A**) and patients with long COVID (**B**) and separately by group for complement regulatory proteins (**C** and **F**, respectively), complement components (**D** and **G**, respectively), and complement activation products (**E** and **H**, respectively). The size of each circle indicates the strength of the correlation, and colors show the intensity of direct (dark blue) or inverse (dark red) correlations, each measured using the Pearson correlation coefficient (*r*).

**Figure 6.**
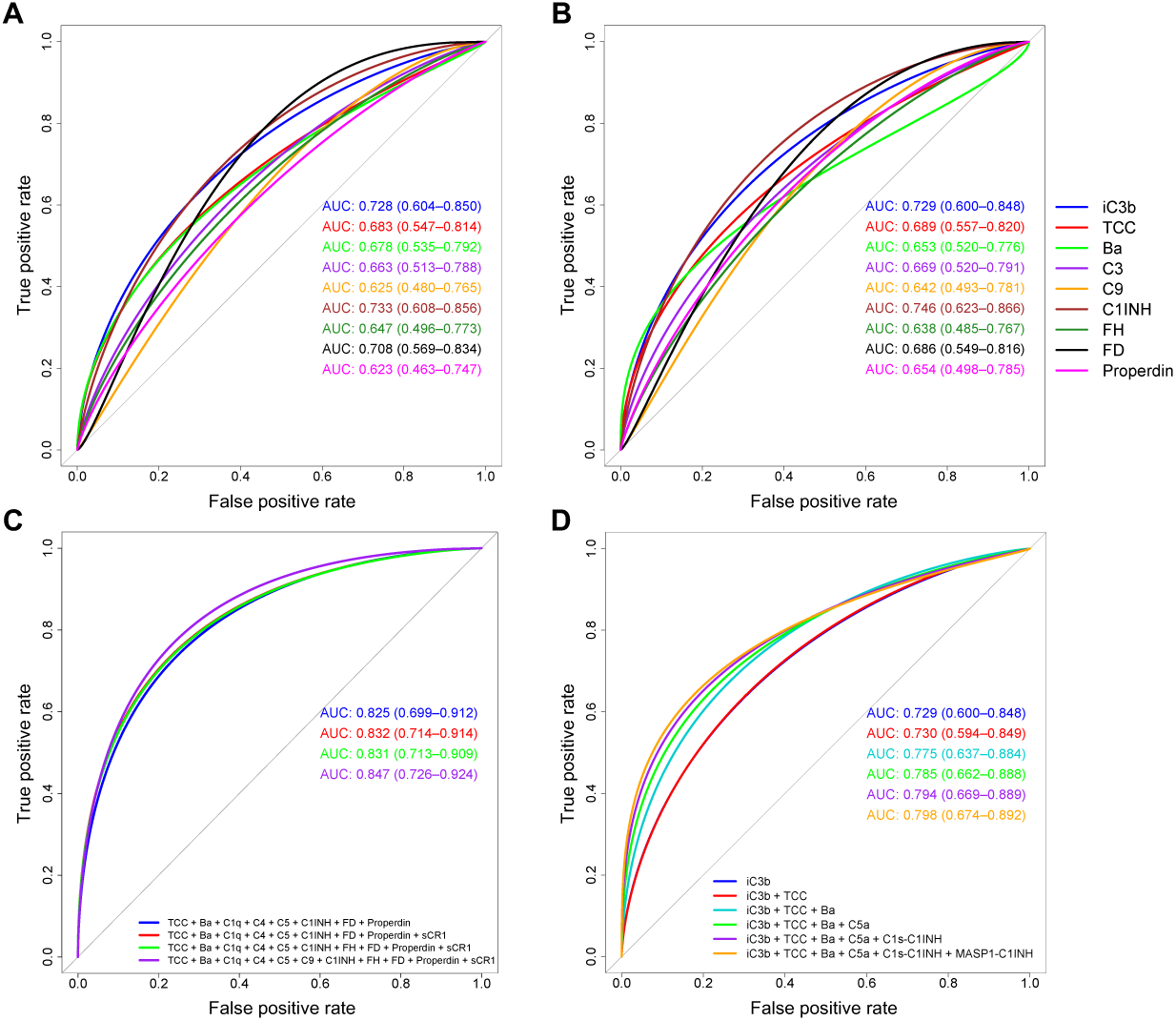
Receiver operator characteristic curves for generalized linear models using complement protein concentrations in plasma to predict long COVID. **(A–D)** Receiver operator curves were generated using multiple generalized linear models (GLMs) for each complement protein. AUC statistics are shown for individual (**A**, **B**) or combined analytes (**C**, **D**) with 95% confidence intervals (generated from 2000 bootstrap replicates) in parentheses for each GLM. Proteins were unadjusted (**A**) or adjusted for age, gender, and BMI (**B**, **C**, and **D**, respectively). Linear predictors were selected according to the results of stepAIC models of adjusted protein concentrations (**C**). The best model was plotted first based on the highest AIC relative to model complexity (TCC + Ba + C1q + C4 + C5 + C1INH + FD + properdin), and then three regressive steps from the stepAIC results were plotted sequentially. Sequential combinations are shown for each of the top ranked AUC statistics from complement activator GLMs (**D**), starting with the highest ranked AUC (iC3b).

The impact of combining the most predictive biomarkers was then tested in ROC analyses via stepAIC model selection. The best model contained TCC, Ba, C1q, C4, C5, C1INH, FD, and properdin, with an overall AUC of 0.825 and an Akaike Information Criterion (AIC) of 141.85 (**Figure 6C**). Sequential regressive steps from the best stepAIC method did not significantly improve either the AUCs or the AICs. In a second analysis focused on activation markers, the combination of iC3b, TCC, Ba, and C5a increased the AUC (0.729 to 0.785) relative to iC3b alone (**Figure 6D**). Inclusion of the other two activation markers, C1s-C1INH and MASP1-C1INH, had no significant impact on the AUC (**Figure 6D**). Forest plots are shown in **Supplementary** Figure 2, and details of all ROC-AUC analyses are provided in **Supplementary Tables 4–8**.

We then conducted a principal component analysis (PCA) (**Supplementary** Figure 3A). Visualization of individual sample contributions to the first two principal components (PCs) revealed considerable overlap between healthy convalescent individuals and patients with long COVID (**Supplementary** Figure 3B). The first two principal components (PCs) explained a total of 35% of the total variance (20.2% for PC1, 14.8% for PC2) (**Supplementary** Figure 3C). C3 made the greatest contribution to PC1 (17.27%), followed by C9 (12.7%) and C5 (11.21%) (**Supplementary** Figure 3D), whereas Ba made the greatest contribution to PC2 (9.74%), followed by C1INH (9.05%) and FHR-125 (9.03%) (**Supplementary** Figure 3E).

Collectively, these data and the associated analyses, especially the GLMs, demonstrate that complement dysregulation is a consistent and predictive feature of long COVID.

## DISCUSSION

Complement dysregulation is a common feature of diverse acute and chronic inflammatory diseases and a major driver of inflammation. There is abundant evidence from biomarker studies to indicate that complement dysregulation involving all activation pathways is ubiquitous during acute infection with SARS-CoV-2, especially in individuals with severe COVID-19.^10–17^ Moreover, admission levels of several complement biomarkers, including the activation fragment Ba, predict outcome in hospitalized patients with acute COVID-19.^10^ Progressive changes in the concentrations of some complement biomarkers have also been reported in the context of severe disease.^10,14–17,19^ In mechanistic terms, complement activation under these circumstances has been variously attributed to direct virus-mediated triggering of the classical, lectin, and/or alternative pathways, activation of the classical pathway via antiviral antibodies, and/or indirect activation via contact with infected cells and/or damaged tissue.^20–24^

These observations have been used to rationalize interventions with complement blocking drugs to mitigate the hyperinflammatory state that characterizes severe COVID-19. Early pilot studies reported positive effects using repurposed existing drugs to inhibit complement activation or the terminal pathway.^25–30^ However, the numerous clinical trials of agents targeting C5, C5a/C5aR, C3, FD, or C1 that followed have been disappointing to date, potentially reflecting a failure to screen for complement dysregulation prior to inclusion, a focus on the most severe cases, and/or treatment late in the disease course.^31^

Persistent inflammation has been implicated as a key component of long COVID. In particular, elevated levels of inflammatory cytokines and the inflammatory markers C-reactive protein and serum amyloid A were detected in a study of symptomatic patients 6–9 months after acute infection with SARS-CoV-2,^7^ and a systematic review compiling data from 22 studies confirmed an association between elevated plasma concentrations of IL-6 and long COVID.^8^ High titers of various autoantibodies, notably those associated with myopathies, vasculitides, and other related conditions, have also been reported in long COVID.^32^ In another study, autoantibodies against inflammatory chemokines were found to be common in the convalescent phase and correlated with better outcomes and a lower risk of developing long COVID.^33^ Innate immune cell activation has also been identified as a driver of lung fibrosis and inflammation in a humanized mouse model of long COVID.^34^

All of these reports point to immune dysregulation with attendant inflammation as critical determinants of long COVID, but none identify an obvious inflammatory trigger or target for therapy.^3^ In this study, we set out to test whether persistent complement dysregulation contributed to the pathophysiology of long COVID. We found that markers of complement activation spanning the classical (C1s-C1INH), alternative (iC3b, Ba), and terminal pathways (C5a, TCC), but not the lectin pathway (MASP1-C1INH), were all significantly elevated in patients with long COVID compared with healthy convalescent controls infected during the same “wave” of SARS-CoV-2. In a previous study, we found that Ba, iC3b, and TCC stood out as markers of disease course and outcome in patients with acute COVID-19.^10^ Moreover, plasma concentrations of iC3b and TCC remained elevated in convalescent samples up to a median of 21 days after discharge, indicating persistent complement dysregulation.^10^ It is particularly notable here that C5a, a leukocyte activator and chemoattractant, and TCC, a proxy for membrane attack complex (MAC) formation, are both potent triggers of inflammation and collaborate across diverse cell types to activate the inflammasome pathway.^35,36^

In further assays, we found that plasma concentrations of many complement components were also relatively elevated in patients with long COVID. These observations could be explained by the fact that C3, C4, C5, and C9 are all positive acute phase reactants, such that measured plasma levels reflect the net effect of consumption caused by complement activation and increased synthesis driven by inflammation.^37^ In contrast, plasma concentrations of C1q, which is not an acute phase reactant, were significantly lower in cases versus controls, likely reflecting uncompensated consumption. Moreover, we found that plasma concentrations of several complement regulators, namely C1INH, FD, properdin, FH, and clusterin, were relatively elevated in patients with long COVID. Elevated levels of C1INH would be expected to limit activation of the classical and lectin pathways, whereas FD, properdin, and FH all regulate the alternative pathway. Specifically, FD mediates the enzymatic cleavage of FB, which is required to form the alternative pathway convertase C3bBb, and properdin stabilizes C3bBb, such that elevated levels of these regulators would be expected to increase activity in the alternative pathway. In contrast, FH is a negative regulator, catalyzing the inactivation of C3bBb. Activation of the alternative pathway nonetheless dominated in patients with long COVID, as highlighted by the elevated plasma concentrations of Ba and iC3b. Clusterin is a multifunctional plasma lipoprotein that inhibits assembly of the MAC. It is notable here that plasma levels of clusterin were previously found to be reduced in severe acute COVID-19.^10^

Data analyses revealed that 9/21 complement analytes measured in our study were predictive of long COVID (AUC > 0.6). The most predictive single biomarker was C1INH, with an AUC of 0.746, and optimal prediction was enabled by a stepAIC-informed combination of biomarkers incorporating Ba, C1q, C4, C5, C1INH, FD, properdin, and TCC. A more clinically tractable combination of just four activation markers, namely Ba, iC3b, C5a, and TCC, yielded an AUC of 0.785. These biomarker sets focus attention on dysregulation of the alternative pathway amplification loop with activation of the downstream terminal pathway, reminiscent of our findings in acute COVID-19.^10^

There are no specific therapies for long COVID. Current treatment approaches focus on symptom relief and multidisciplinary rehabilitation.^3^ A handful of clinical trials are in progress using drugs that target cardiac damage (ivabradine), fibrotic lung injury (pirfenidone, inhaled interferon-1), and inflammation (leronlimab) induced by acute infection with SAR-CoV-2.^38^ However, the patients in our cohort exhibited no concomitant symptom-driving pathology, and allied tests for underlying organ dysfunction universally fell within the normal range. Accordingly, our data suggest that complement dysregulation and the associated inflammatory response are viable targets for therapeutic interventions designed to ameliorate symptoms and break the pathogenic cycle of disease. There are now many complement inhibitors in clinical use that could be repurposed to treat long COVID. Our findings suggest that the preferred target is the alternative pathway, which can be suppressed by drugs such as pegcetacoplan (targeting C3), iptacopan (targeting FB), and vemircopan (targeting FD).^39^ On this basis, we propose that pilot trials are now warranted to test the efficacy of such drugs under close clinical supervision, even for a relatively short period of time, with the aim of disrupting the proinflammatory cycle and restoring a normal pattern of homeostasis in patients with long COVID.

### Limitations of the study

Our analysis was cross-sectional in nature, precluding retrospective associations with complement activation and clinical severity at the time of infection, and our study population was predominantly Caucasian. Moreover, the selection of biological confounders as adjustment variables was restricted to age, gender, and BMI, which did not necessarily account for all of the observed variance. In addition, statistical power was potentially limited by sample size, given the lack of significance for several complement proteins included in the GLMs. Sample size also precluded meaningful segregation based on dominant symptoms, symptom clusters, or functional disability. It is further notable that our study was not designed to identify the precise triggers of complement activation in long COVID.

## METHODS

### Study overview

EDTA plasma samples were collected from age/ethnicity/gender/infection/vaccine-matched healthy convalescent individuals (controls, *n* = 79) and patients with long COVID (cases, *n* = 166). All participants had a clinical history of acute COVID-19 and direct evidence of infection with SARS-CoV-2. Cases were diagnosed according to the National Institute for Health and Care Excellence (NICE) guideline NG188 (https://www.nice.org.uk/guidance/ng188). Eligible patients were men and non-pregnant women over the age of 18 years with no alternative underlying disease and symptoms that persisted for at least 12 weeks after the initial diagnosis of acute COVID-19. In 46.8% of controls and 54.8% of cases, the index infection occurred >2 years prior to sample acquisition, which was limited to a time window between February and October 2022. Symptoms were scored individually using a numeric self-rating scale from 0 (no symptom) to 10 (worst possible symptom). Overall general health was scored similarly on an inverse scale from 0 (worst possible) to 10 (best possible). All participants provided written informed consent in accordance with the principles of the Declaration of Helsinki. Study approval was granted by the Cardiff University School of Medicine Research Ethics Committee (21/55) and by the Health Research Authority and Health and Care Research Wales (20/NW/0240). Cohort demographics, symptomatology, and other key features are summarized in **Supplementary Table 1**.

### Immunoassays

EDTA blood samples were kept on ice immediately after acquisition. Plasma was separated promptly and stored in aliquots at −80°C. Complement components (C1q, C3, C4, FB, C5, and C9), regulators (C1INH, FH, FHR125, FHR4, FI, FD, properdin, sCR1, and clusterin), and activation products (C1s-C1INH, MASP1-C1INH, Ba, iC3b, C5a, and TCC) were quantified using established in-house or commercial ELISAs. For in-house assays, 96-well MaxiSorp plates (Nunc) were coated with the relevant affinity-purified capture antibody overnight at 4°C, blocked with 2% bovine serum albumin (BSA) or 1% non-fat-dried milk (NFM) in phosphate-buffered saline containing 0.1% Tween 20 (PBST, Sigma-Aldrich) for 1 h at room temperature (RT), and washed with PBST. Plasma samples and purified proteins standards optimally diluted in 0.2% BSA or 0.1% NFM in PBST were then added in duplicate (50 µl/well) and incubated for 1 h at RT. After a further wash with PBST, the relevant detection antibody (biotinylated, unlabeled, or labeled with horseradish peroxidase [HRP] in-house using commercially available kits, Thermo Fisher Scientific) was added for 1 h at RT. The plates were then washed with PBST, incubated with streptavidin-HRP or a secondary antibody (HRP-labeled anti-IgG or HRP-labeled anti-IgM) as appropriate, washed again with PBST, and developed using O-phenylenediamine dihydrochloride (SIGMAFAST OPD, Sigma-Aldrich) or tetramethylbenzidine (Thermo Fisher Scientific). Reactions were stopped using 5% sulphuric acid, and absorbances were read at 450 nm or 492 nm. Standard curves were fitted using a non-linear regression model. Sample protein concentrations for each analyte were automatically calculated with reference to the corresponding curve using Prism version 9.5.0 (GraphPad). Plasma dilutions for each biomarker were selected to fall within the linear portion of the log standard curve. All assays passed quality control tests, including evaluations of sensitivity, reproducibility, and coefficients of variation within and across assays (each <10%). Individual assay details are provided in **Supplementary Table 2**.

### Anti-RBD IgG assay

Antibodies against the SARS-CoV-2 spike protein RBD were detected using a direct ELISA.^40^ In brief, 96-well MaxiSorp plates (Nunc) were coated overnight with recombinant RBD protein (2 µg/ml) at 4°C, blocked with 3% NFM in PBST for 1 h at RT, and washed with PBST. Plasma samples diluted 1 in 50 in 1% NFM in PBST were then added in duplicate and incubated for 2 h at RT. After a further wash with PBST, donkey anti-human IgG F(ab’)2-HRP (Jackson ImmunoResearch) was added for 1 h at RT. The plates were then washed again with PBST and developed using O-phenylenediamine dihydrochloride (SIGMAFAST OPD, Sigma-Aldrich). Absorbance was measured at 492 nm.

### Haemolytic assay

Classical pathway haemolytic activity was measured using sheep erythrocytes sensitized with rabbit anti-sheep erythrocyte antiserum (Siemens Amboceptor, Cruinn Diagnostics). Sensitized sheep erythrocytes (EA) were diluted to 2% in HEPES-buffered saline (HBS) comprising 0.01 M HEPES, 0.15 M NaCl, 2 mM CaCl_2_, and 1 mM MgCl_2_. Plasma samples were diluted sequentially in HBS and then added in duplicate (50 µl/well) with EA (50 µl/well) and HBS (50 µl/well) to a 96-well tissue-culture plate (Nunc). Assays were incubated for 30 min at 37°C. Intact cells were pelleted via centrifugation. Haemolysis was measured by reading absorbance in the supernatant at 540 nm. Percent haemolysis in each experimental well was calculated relative to the negative (0% lysis) and positive control (100% lysis) wells to determine the 50% classical haemolytic dilution (CH_50_).

### Statistics

Categorical variables were assessed using the chi-square test, group means were compared using unpaired t-tests, and relationships among variables were evaluated using Pearson’s correlation. Significance was assigned at p < 0.05. All statistical analyses were performed using Prism version 9.5.0 (GraphPad).

### Receiver operating characteristic analysis

A series of GLMs using different combinations of protein measurement data with varying complexity were constructed using the base *stats* package in R, with a binomial model for error distribution and specified link function. Data were randomly split 70/30 into “training” and “test” sets to prevent overfitting and stratified to maintain case/control proportions, and “test” data were reported as AUCs. GLMs containing three major confounders (age, sex, and BMI) were compared to GLMs containing the same confounders and each complement protein using the Delong test to show the effects of each analyte on the resultant AUCs. Protein levels were adjusted for age and sex and standardized to a mean of 0 and a standard deviation of 1 to maintain equal contributions of each protein to the analyses and prevent bias arising from proteins with wider ranges, and analyses of unadjusted protein levels were used for comparison. A stepAIC model was run to inform the best features to keep in the final model via iterative analysis of AICs. Models with fewer protein measurements were favored to promote the general applicability of our approach. Sequential combinations adjusted complement proteins were also included in a series of GLMs. The order of inclusion was informed by the corresponding ranked AUCs. Complement protein concentration distributions were compared using the Wilcoxon-Mann-Whitney test, and models were compared using ROC curves, with 95% confidence intervals calculated via the default “bootstrap” method across 2000 replicates for each AUC.

### Principal component analysis

The contribution of each measured protein to the overall variance of individual protein levels and samples was assessed via PCA using the base *stats* package in R. Data were scaled and adjusted as described above. Each principal component and the contribution of each protein to the top two principal components, together with the contribution of individual samples to the overall variance, were visualized using *factoextra* with *ggplot2* and *ggpubr* in R.

### Data visualization

Data were visualized as cluster plots and histograms using the *ggplot2* package in R. Correlation plots showing the Pearson correlation coefficient (*r*) for each complement protein quantified in controls versus cases were created using the base *stats* package in R and visualized using the *corrplot* package in R.

## Data Availability

All data produced in the present work are shown in the manuscript. Raw data files are available upon reasonable request from the lead contact.

## ACKNOWLEDGEMENTS

We thank all participants for their enthusiastic contributions to this study. This work was funded by the National Institute for Health Research (COV-LT2-0041), the PolyBio Research Foundation, and the UK Dementia Research Institute. Additional support was provided via an Alzheimer’s Research UK Race Against Dementia Fellowship Award (W.M.Z.). We also thank Lisa Hurler, Erika Kajdácsi, László Cervenak, and Zoltán Prohászka (Department of Medicine and Haematology, Semmelweis University, Budapest, Hungary) for developing the C1s/C1-INH and MASP-1/C1-INH complex ELISAs, which were kindly donated by Loek Willems (Hycult Biotech, Eindhoven, Netherlands).

## AUTHOR CONTRIBUTIONS

K.B., S.B.K.K., K.L., K.L.M., S.A.J., E.M., E.J.M.T., and W.M.Z. performed experiments and/or provided reagents/resources and/or analyzed data; H.E.D., D.A.P., B.P.M., and W.M.Z. conceived the project, led the study, supervised the work, and wrote the paper. All authors approved the final draft of the manuscript and concurred with the decision to submit for publication.

