## Supplemental Information File for "Complement dysregulation is a predictive and therapeutically amenable feature of long COVID"

**Supplementary Table 1.** Demographics, symptomatology, and other key features of healthy convalescent individuals and patients with long COVID.

| Characteristic | Controls (*n* = 79) | Long COVID (*n* = 166) |
|---|---|---|
| **Age (years), median (range)** | 45 (21–82) | 47 (20–83) |
| **Female** | 62 (78.5%) | 127 (76.5%) |
| **Ethnicity/race** | | |
| White | 67 (84.8%) | 146 (88.0%) |
| Black | 0 (0.0%) | 3 (1.8%) |
| Asian | 8 (10.2%) | 8 (4.8%) |
| Mixed ethnicity | 2 (2.5%) | 2 (1.2%) |
| Other | 2 (2.5%) | 7 (4.2%) |
| **Body mass index >30kg/m$^2$** | 26/75 (34.6%) | 79/162 (48.8%) |
| **Coexisting conditions** | | |
| Yes | 14 (17.7%) | 55 (33.1%) |
| No | 65 (82.3%) | 111 (66.9%) |
| **Employment status** | | |
| *Pre-COVID* | | |
| Employed | 53 (91.4%) | 139 (89.1%) |
| Employed, altered duties | 0 (0.0%) | 0 (0.0%) |
| Employed, sick leave | 0 (0.0%) | 0 (0.0%) |
| Unemployed | 0 (0.0%) | 4 (2.6%) |
| Retired | 2 (3.4%) | 11 (7.0%) |
| Student | 3 (5.2%) | 2 (1.3%) |
| *Post-COVID* | | |
| Employed | 54 (93.2%) | 68 (43.6%) |
| Employed, altered duties | 0 (0.0%) | 28 (17.9%) |
| Employed, sick leave | 0 (0.0%) | 33 (21.2%) |
| Unemployed | 0 (0.0%) | 11 (7.1%) |
| Retired | 2 (3.4%) | 13 (8.3%) |
| Student | 2 (3.4%) | 3 (1.9%) |
| **Date of infection with SARS-CoV-2** | | |
| March 2020 to August 2020 | 14 (17.7%) | 34 (20.5%) |
| September 2020 to June 2021 | 23 (29.1%) | 57 (34.3%) |
| July 2021 to October 2022 | 42 (53.2%) | 75 (45.2%) |
| **COVID vaccination status, median (IQR)** | | |
| Number of vaccinations before infection | 3 (0–3) | 2 (0–2) |
| Total number of vaccinations | 3 (3–3) | 3 (3–3) |
| **Symptoms**[1] | | |
| Breathlessness | 0 (0–0) | 4 (2–6) |
| Fatigue | 0 (0–2) | 6 (4–8) |
| Musculoskeletal | 0 (0–0) | 3 (0–6) |
| Pain | 0 (0–0) | 4 (2–6) |
| Neuropsychiatric | 0 (0–0) | 3 (1–5) |
| **Ability to maintain self-care** | 0 (0–0) | 0 (0–3) |
| **Ability to maintain daily tasks** | 0 (0–0) | 6 (3–8) |
| **Overall general health**[2] | 0 (0 to −1) | −4 (−2 to −6) |

[1]Difference in numeric rating scale score (0 = no symptom, 10 = worst possible symptom) before versus after COVID-19 (median ± IQR). [2]Difference in numeric rating scale score (0 = worst possible, 10 = best possible) before versus after COVID-19 (median ± IQR).

**Supplementary Table 2.** Summary of optimized in-house ELISAs.

| Assay | Capture antibody | Detection antibody | Detection range | Dilution factor |
|---|---|---|---|---|
| *Activation products* | | | | |
| Ba | Anti-Ba (clone D22/3, Hycult) | Anti-FB–biotin (clone P21/15, Hycult) | 8.5–550 ng/ml | 1:4 |
| iC3b | Anti-iC3b (clone 9) | Anti-C3c–HRP (mAb bH6, Hycult) | 23–1500 ng/ml | 1:40 |
| TCC | Anti-TCC (clone aE11, Hycult) | Anti-C8–biotin (mAb E2) | 39–2500 ng/ml | 1:40 |
| *Components* | | | | |
| C1q | Anti-C1q (clone 9H10) | Anti-C1q (rabbit polyclonal) | 23–1500 ng/ml | 1:16,000 |
| C4 | Anti-C4 (rabbit polyclonal) | Anti-C4–HRP (rabbit polyclonal) | 15–1000 ng/ml | 1:4000 |
| C3 | Anti-C3/C3a (clone 2898, Hycult) | Anti-C3d–HRP (clone 3, Hycult) | 31–2000 ng/ml | 1:2000 |
| FB | Anti-FB (clone JC1) | Anti-FB (goat polyclonal, CompTech) | 15–1000 ng/ml | 1:3000 |
| C5 | Anti-C5 (clone 10B6) | Anti-C5 (clone 4G2) | 39–2500 ng/ml | 1:2000 |
| C9 | Anti-C9 (clone B7) | Anti-C9–biotin (rabbit polyclonal) | 3–200 ng/ml | 1:4000 |
| *Regulators* | | | | |
| C1INH | Anti-C1INH (rabbit polyclonal) | Anti-C1INH–HRP (rabbit polyclonal) | 1–125 ng/ml | 1:4000 |
| FH | Anti-FH (clone OX24) | Anti-FH–HRP (clone 35H9) | 15–1000 ng/ml | 1:4000 |
| FHR125 | Anti-FHR125 (clone MBI125) | Anti-FH (clone 35H9) | 156–10,000 ng/ml | 1:100 |
| FHR4 | Anti-FHR4 (clone 4E9) | Anti-FHR4–HRP (clone 150) | 15–1000 ng/ml | 1:100 |
| sCR1 | Anti-sCR1 (rabbit polyclonal) | Anti-sCR1–HRP (mAb MBI35) | 0.4–25 ng/ml | 1:2 |
| Properdin | Anti-properdin (clone 9.3.4, Hycult) | Anti-properdin–HRP (clone 2.9, Hycult) | 6–400 ng/ml | 1:1600 |
| FD | Anti-FD (rabbit polyclonal) | Anti-FD–HRP (clone D10/4, Hycult) | 15–1000 ng/ml | 1:100 |
| FI | Anti-FI (clone 7B5) | Anti-FI (rabbit polyclonal) | 15–1000 ng/ml | 1:500 |
| Clusterin | Anti-clusterin (clone 2D5) | Anti-clusterin–HRP (clone 4C7) | 31–2000 ng/ml | 1:1000 |

All antibodies were prepared in-house unless stated otherwise. All standards were prepared in-house, except for iC3b, Ba, properdin, FD, and FB (all from CompTech), and C1INH (Cinryze, Shire Pharma). C1s-C1INH was assayed using a Human C1s/C1-INH Complex ELISA (Hycult Biotech), C5a was assayed using a Human Complement Component C5a Duoset ELISA (R&D Systems), and MASP1-C1INH was assayed using a Human MASP-1/C1-INH Complex ELISA (Hycult Biotech). TCC, terminal complement complex; FB, factor B; C1INH, C1 inhibitor; FH, factor H; FHR125, FH-related proteins 1, 2, and 5; FHR4, FH-related protein 4; sCR1, soluble complement receptor type 1; FD, factor D; FI, factor I; HRP, horseradish peroxidase.

**Supplementary Table 3.** Statistics for comparing the distributions of complement protein concentrations in plasma in healthy convalescent individuals and patients with long COVID.

| Protein | Control Median | Control IQR | Long COVID Median | Long COVID IQR | P value |
|---|---|---|---|---|---|
| *Activation products* | | | | | |
| C1s-C1INH µg/ml | 1.114 | 0.470 | 1.191 | 0.602 | **0.0237** |
| MASP1-C1INH ng/ml | 51.998 | 37.595 | 49.706 | 52.723 | 0.644 |
| Ba µg/ml | 0.191 | 0.149 | 0.221 | 0.249 | **0.00889** |
| iC3b µg/ml | 11.904 | 7.132 | 17.089 | 13.487 | **2.29E−08** |
| C5a ng/ml | 4.533 | 3.142 | 5.275 | 3.411 | 0.0855 |
| TCC µg/ml | 3.426 | 2.436 | 5.301 | 3.103 | **2.34E−08** |
| *Components* | | | | | |
| C1q µg/ml | 117.916 | 137.751 | 108.310 | 121.844 | **0.023** |
| C4 µg/ml | 426.654 | 214.485 | 468.643 | 189.476 | 0.836 |
| C3 µg/ml | 836.095 | 191.517 | 872.514 | 205.933 | **0.0408** |
| FB µg/ml | 245.815 | 117.479 | 250.730 | 137.235 | 0.637 |
| C5 µg/ml | 175.114 | 53.613 | 191.596 | 65.589 | **0.00314** |
| C9 µg/ml | 83.639 | 33.869 | 89.597 | 36.443 | **0.0163** |
| *Regulators* | | | | | |
| C1INH µg/ml | 96.992 | 40.394 | 113.515 | 45.481 | **0.000621** |
| FH µg/ml | 233.574 | 118.639 | 250.472 | 94.885 | 0.0677 |
| FHR125 µg/ml | 79.938 | 82.719 | 85.551 | 87.266 | 0.471 |
| FHR4 µg/ml | 3.724 | 1.907 | 3.573 | 1.959 | 0.735 |
| sCR1 ng/ml | 6.183 | 1.956 | 6.202 | 2.075 | 0.327 |
| Properdin µg/ml | 2.931 | 0.942 | 3.425 | 1.391 | **7.36E−05** |
| FD µg/ml | 0.618 | 0.294 | 0.844 | 0.370 | **2.88E−07** |
| FI µg/ml | 53.164 | 15.724 | 54.093 | 18.685 | 0.539 |
| Clusterin µg/ml | 131.67 | 25.733 | 137.644 | 66.457 | 0.228 |

P values were calculated using the Wilcoxon-Mann-Whitney test. Significant differences are highlighted in bold font. C1INH, C1 inhibitor; MASP1, mannose-associated serine protease 1; TCC, terminal complement complex; FB, factor B; FH, factor H; FHR125, FH-related proteins 1, 2, and 5; FHR4, FH-related protein 4; sCR1, soluble complement receptor type 1; FD, factor D; FI, factor I.

**Supplementary Table 4.** Statistics for generalized linear models using unadjusted complement protein concentrations in plasma to predict long COVID.

| Protein | AUC | CI | Z | SE | P value |
|---|---|---|---|---|---|
| iC3b | 0.744 | 0.617–0.859 | 2.388 | 0.228 | **0.0169** |
| TCC | 0.687 | 0.547–0.814 | 3.765 | 0.262 | **0.000166** |
| Ba | 0.679 | 0.542–0.802 | 2.065 | 0.357 | **0.0389** |
| C1q | 0.452 | 0.312–0.601 | −1.342 | 0.176 | 0.18 |
| C3 | 0.667 | 0.525–0.797 | 1.105 | 0.185 | 0.269 |
| C4 | 0.536 | 0.369–0.694 | −0.956 | 0.178 | 0.339 |
| C5 | 0.512 | 0.352–0.671 | 3.203 | 0.219 | **0.00136** |
| C9 | 0.625 | 0.47–0.768 | 2.278 | 0.193 | **0.0227** |
| C1INH | 0.744 | 0.614–0.859 | 1.741 | 0.184 | **0.0817** |
| FH | 0.65 | 0.507–0.785 | 1.15 | 0.186 | 0.25 |
| FHR4 | 0.485 | 0.337–0.627 | −0.509 | 0.17 | 0.61 |
| FHR125 | 0.484 | 0.341–0.632 | 0.306 | 0.185 | 0.759 |
| FI | 0.521 | 0.368–0.671 | 1.091 | 0.167 | 0.275 |
| Clusterin | 0.554 | 0.422–0.683 | 1.264 | 0.201 | 0.206 |
| FD | 0.707 | 0.566–0.843 | 2.392 | 0.254 | **0.0167** |
| Properdin | 0.618 | 0.467–0.752 | 3.157 | 0.314 | **0.0016** |
| FB | 0.528 | 0.356–0.688 | 0.526 | 0.193 | 0.599 |
| sCR1 | 0.626 | 0.487–0.759 | −0.604 | 0.161 | 0.546 |
| C1s-C1INH | 0.567 | 0.418–0.719 | 1.567 | 0.186 | 0.117 |
| MASP1-C1INH | 0.553 | 0.399–0.699 | 0.825 | 0.211 | 0.409 |
| C5a | 0.544 | 0.391–0.694 | 1.707 | 0.384 | 0.0879 |

AUC, area under the curve; CI, 95% confidence interval (generated from 2000 bootstrap replicates); Z, Z score; SE, standard error. Significant differences are highlighted in bold font. TCC, terminal complement complex; C1INH, C1 inhibitor; FH, factor H; FHR4, FH-related protein 4; FHR125, FH-related proteins 1, 2, and 5; FI, factor I; FD, factor D; FB, factor B; sCR1, soluble complement receptor type 1; MASP1, mannose-associated serine protease 1.

**Supplementary Table 5.** Statistics for generalized linear models using complement protein concentrations in plasma adjusted for age, gender, and BMI to predict long COVID.

| **Protein** | **AUC** | **CI** | **Z** | **SE** | **P value** |
|---|---|---|---|---|---|
| iC3b | 0.729 | 0.6–0.848 | 2.325 | 0.226 | **0.0201** |
| TCC | 0.689 | 0.557–0.82 | 3.669 | 0.282 | **0.000244** |
| Ba | 0.653 | 0.52–0.776 | 2.182 | 0.336 | **0.0291** |
| C1q | 0.434 | 0.322–0.613 | −1.191 | 0.174 | 0.234 |
| C3 | 0.669 | 0.52–0.791 | 0.905 | 0.183 | 0.365 |
| C4 | 0.507 | 0.364–0.674 | −1.203 | 0.17 | 0.229 |
| C5 | 0.509 | 0.353–0.659 | 3.052 | 0.224 | **0.00228** |
| C9 | 0.642 | 0.493–0.781 | 2.051 | 0.19 | **0.0403** |
| C1INH | 0.746 | 0.623–0.866 | 1.729 | 0.183 | 0.0839 |
| FH | 0.638 | 0.485–0.767 | 1.078 | 0.177 | 0.281 |
| FHR4 | 0.522 | 0.387–0.674 | −0.824 | 0.172 | 0.41 |
| FHR125 | 0.446 | 0.308–0.6 | 0.467 | 0.179 | 0.64 |
| FI | 0.528 | 0.397–0.679 | 0.669 | 0.176 | 0.504 |
| Clusterin | 0.591 | 0.459–0.713 | 1.363 | 0.19 | 0.173 |
| FD | 0.686 | 0.549–0.816 | 2.329 | 0.23 | **0.0198** |
| Properdin | 0.654 | 0.498–0.785 | 3.161 | 0.292 | **0.00157** |
| FB | 0.518 | 0.35–0.678 | 0.408 | 0.177 | 0.684 |
| sCR1 | 0.599 | 0.449–0.732 | −0.65 | 0.169 | 0.516 |
| C1s-C1INH | 0.575 | 0.431–0.726 | 1.419 | 0.182 | 0.156 |
| MASP1-C1INH | 0.555 | 0.396–0.692 | 0.835 | 0.185 | 0.404 |
| C5a | 0.634 | 0.491–0.762 | 1.849 | 0.384 | 0.0644 |

AUC, area under the curve; CI, 95% confidence interval (generated from 2000 bootstrap replicates); Z, Z score; SE, standard error. Significant differences are highlighted in bold font. TCC, terminal complement complex; C1INH, C1 inhibitor; FH, factor H; FHR4, FH-related protein 4; FHR125, FH-related proteins 1, 2, and 5; FI, factor I; FD, factor D; FB, factor B; sCR1, soluble complement receptor type 1; MASP1, mannose-associated serine protease 1.

**Supplementary Table 6.** Statistics for stepAIC-informed generalized linear models using complement protein concentrations in plasma adjusted for age and gender to predict long COVID.

| Model | AUC | AIC | CI | Z | SE | P value |
|---|---|---|---|---|---|---|
| TCC + Ba + C1q + C4 + C5 + C1INH + FD + properdin | 0.825 | 141.85 | 0.699–0.912 | 5.16 | 0.339 | 2.46E−07 |
| TCC + Ba + C1q + C4 + C5 + C1INH + FD + properdin + sCR1 | 0.832 | 142.62 | 0.714–0.914 | 5.196 | 0.343 | 2.03E−07 |
| TCC + Ba + C1q + C4 + C5 + C1INH + FH + FD + properdin + sCR1 | 0.831 | 143.43 | 0.713–0.909 | 5.207 | 0.352 | 1.91E−07 |
| TCC+ Ba + C1q + C4 + C5 + C9 + C1INH + FH + FD + properdin + sCR1 | 0.847 | 144.45 | 0.726–0.924 | 5.163 | 0.351 | 2.42E−07 |

AUC, area under the curve; AIC, Akaike Information Criterion; CI, 95% confidence interval (generated from 2000 bootstrap replicates); Z, Z score; SE, standard error. TCC, terminal complement complex; C1INH, C1 inhibitor; FD, factor D; sCR1, soluble complement receptor type 1; FH, factor H.

**Supplementary Table 7.** Statistics for sequential activation generalized linear models using complement protein concentrations in plasma adjusted for age and gender to predict long COVID.

| **Proteins** | **AUC** | **CI** | **Z** | **SE** | **P value** |
|---|---|---|---|---|---|
| iC3b | 0.729 | 0.6–0.848 | 4.796 | 0.183 | 1.62E−06 |
| iC3b + TCC | 0.73 | 0.594–0.849 | 4.965 | 0.21 | 6.87E−07 |
| iC3b + TCC + Ba | 0.775 | 0.637–0.884 | 4.991 | 0.234 | 6.02E−07 |
| iC3b + TCC + Ba + C5a | 0.785 | 0.662–0.888 | 4.921 | 0.248 | 8.6E−07 |
| iC3b + TCC + Ba + C5a + C1s-C1INH | 0.794 | 0.669–0.889 | 4.945 | 0.254 | 7.61E−07 |
| iC3b + TCC + Ba + C5a + C1s-C1INH + MASP1-C1INH | 0.798 | 0.674–0.892 | 4.931 | 0.257 | 8.19E−07 |

AUC, area under the curve; CI, 95% confidence interval (generated from 2000 bootstrap replicates); Z, Z score; SE, standard error. Activators of complement proteins were chosen based on the order of individual AUCs, starting with the highest ranked protein (iC3b). TCC, terminal complement complex; C1INH, C1 inhibitor; MASP1, mannose-associated serine protease 1.

**Supplementary Table 8.** Statistics for main three confounders (age, gender, and BMI) in generalized linear models using complement biomarker concentrations in plasma to predict long COVID.

| Model | AUC | CI | Z | SE | P value | CI DeLong | Z DeLong | P value DeLong |
|---|---|---|---|---|---|---|---|---|
| Age + gender + BMI | 0.491 | 0.332–0.658 | 1.077 | 0.014 | 0.281 | NA | NA | NA |
| + iC3b | 0.723 | 0.587–0.846 | 2.34 | 0.228 | 0.0193 | −0.0947 | −3.31 | 0.000928 |
| + TCC | 0.708 | 0.571–0.836 | 3.647 | 0.264 | 0.000265 | −0.042 | −2.43 | 0.0151 |
| + Ba | 0.636 | 0.503–0.756 | 2.15 | 0.369 | 0.0316 | −0.0173 | −2.23 | 0.0261 |
| + C1q | 0.477 | 0.326–0.624 | −1.18 | 0.179 | 0.238 | 0.302 | 0.0967 | 0.923 |
| + C3 | 0.599 | 0.441–0.752 | 0.901 | 0.188 | 0.368 | −0.0264 | −2.6 | 0.00932 |
| + C4 | 0.48 | 0.333–0.617 | −1.247 | 0.184 | 0.213 | 0.263 | 0.0866 | 0.931 |
| + C5 | 0.527 | 0.365–0.686 | 3.066 | 0.219 | 0.00217 | 0.242 | −0.25 | 0.802 |
| + C9 | 0.596 | 0.444–0.744 | 2.064 | 0.201 | 0.039 | 0.0439 | −1.38 | 0.168 |
| + C1INH | 0.715 | 0.567–0.843 | 1.73 | 0.185 | 0.0837 | −0.0968 | −3.45 | 0.000553 |
| + FH | 0.621 | 0.476–0.769 | 1.068 | 0.188 | 0.286 | −0.00639 | −2.06 | 0.0392 |
| + FHR4 | 0.499 | 0.339–0.659 | −0.84 | 0.177 | 0.401 | 0.0724 | −0.197 | 0.844 |
| + FHR125 | 0.551 | 0.395–0.701 | 0.472 | 0.191 | 0.637 | −0.0136 | −2.54 | 0.0112 |
| + FI | 0.515 | 0.358–0.666 | 0.669 | 0.177 | 0.503 | 0.0454 | −0.665 | 0.506 |
| + clusterin | 0.556 | 0.42–0.689 | 1.357 | 0.209 | 0.175 | 0.0493 | −1.11 | 0.266 |
| + FD | 0.639 | 0.503–0.776 | 2.329 | 0.256 | 0.0198 | 0.0117 | −1.82 | 0.0693 |
| + properdin | 0.624 | 0.468–0.761 | 3.153 | 0.323 | 0.00162 | 0.0239 | −1.66 | 0.0968 |
| + FB | 0.483 | 0.329–0.645 | 0.397 | 0.195 | 0.691 | 0.0784 | 0.226 | 0.821 |
| + sCR1 | 0.547 | 0.4–0.697 | −0.692 | 0.169 | 0.489 | 0.0199 | −1.44 | 0.149 |
| + C1s-C1INH | 0.548 | 0.411–0.687 | 1.415 | 0.186 | 0.157 | 0.0751 | −0.843 | 0.399 |
| + MASP1-C1INH | 0.475 | 0.325–0.624 | 0.832 | 0.211 | 0.406 | 0.307 | 0.109 | 0.913 |
| + C5a | 0.575 | 0.42–0.723 | 1.757 | 0.4 | 0.0789 | 0.022 | −1.55 | 0.12 |

AUC, area under the curve; CI, 95% confidence interval (generated from 2000 bootstrap replicates); Z, Z score; SE, standard error. DeLong statistics were used to compare AUCs. TCC, terminal complement complex; C1INH, C1 inhibitor; FH, factor H; FHR4, FH-related protein 4; FHR125, FH-related proteins

1, 2, and 5; FI, factor I; FD, factor D; FB, factor B; sCR1, soluble complement receptor type 1; MASP1, mannose-associated serine protease 1.

#### SUPPLEMENTARY FIGURE LEGENDS

**Supplementary Figure 1. Plasma concentrations of other complement proteins, anti-SARS-CoV-2 spike protein RBD antibody titers, and plasma haemolytic activity in healthy convalescent individuals and patients with long COVID.** (**A**–**D**) Dot plots showing plasma concentrations of FHR4 (**A**) and sCR1 (**B**), anti-SARS-CoV-2 spike protein RBD antibody titers (**C**), and plasma haemolytic activity ($CH_{50}$) (**D**) in healthy convalescent individuals ($n$ = 79) and patients with long COVID ($n$ = 164–166). Horizontal bars represent mean values. Significance was assessed using an unpaired t-test.

**Supplementary Figure 2. Forest plot of regression coefficients (estimated values) from generalized linear models using complement protein concentrations in plasma to predict long COVID.** The direction and strength of effect for each complement protein is shown for healthy convalescent individuals and patients with long COVID. Proteins with positive coefficients are shown in red, and proteins that did not achieve significance are shown in grey. Error bars indicate the standard error for each regression coefficient. *$p < 0.05$, **$p < 0.01$, ***$p < 0.001$.

**Supplementary Figure 3. Principal component analysis of complement analytes measured in this study.** (**A**) Variable correlation plot combining cosine$^2$ (cos2) scores showing the quality of representation for each principal component (PC). (**B**) Plot of individuals showing the quality of representation of each sample colored by clinical phenotype. Positively correlated variables cluster together, and negatively correlated variables are positioned on opposite sides of the plot origin (**A**, **B**). (**C**) Scree plot showing the percent contribution of variance for each PC. (**D**, **E**) Bar plots showing the percent contribution of each complement protein to PC1 (**D**) and PC2 (**E**).

### Supplementary Figure 1

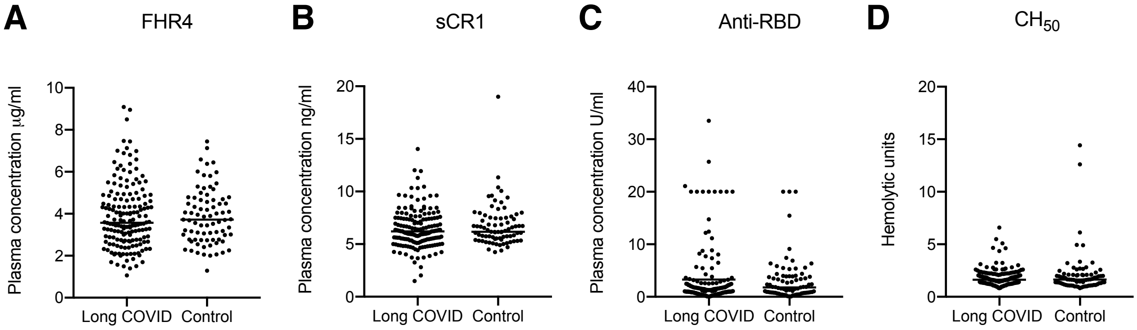

#### Supplementary Figure 2

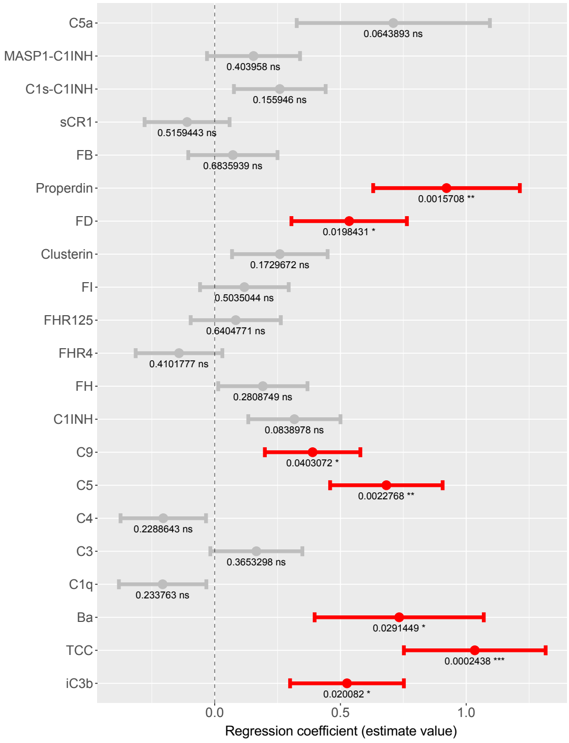

**Supplementary Figure 3**

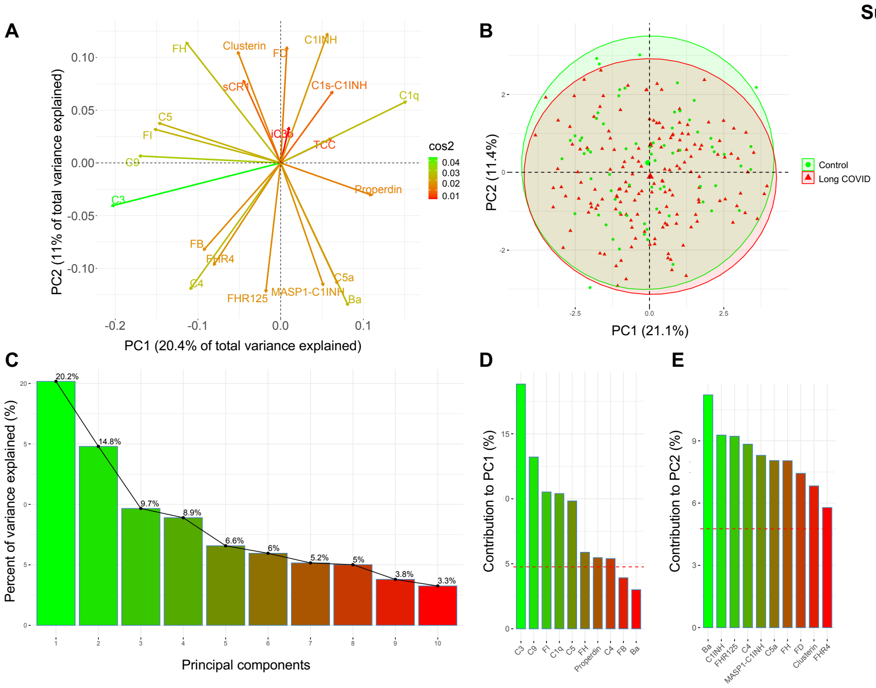
